# A hypothetical intervention to reduce inequities in anxiety for Multiracial people: simulating an intervention on childhood adversity

**DOI:** 10.1101/2023.06.04.23290940

**Authors:** Tracy Lam-Hine, Patrick T. Bradshaw, Amani M. Allen, Michael Omi, Corinne A. Riddell

## Abstract

Multiracial people report higher mean Adverse Childhood Experiences (ACEs) scores and prevalence of anxiety than other racial groups. Studies using statistical interactions to estimate racial differences in ACEs-anxiety associations do not show stronger associations for Multiracial people. Using data from Waves 1 (1995-97) through 4 (2008-09) of the National Longitudinal Study of Adolescent to Adult Health (Add Health), we simulated a stochastic intervention over 1,000 resampled datasets to estimate the race-specific cases averted per 1,000 of anxiety if all racial groups had the same exposure distribution of ACEs as Whites. Simulated cases averted were greatest for the Multiracial group, (median = -4.17 cases per 1,000, 95% CI: -7.42, -1.86). The model also predicted smaller risk reductions for Black participants (-0.76, 95% CI: -1.53, -0.19). CIs around estimates for other racial groups included the null. An intervention to reduce racial disparities in exposure to ACEs could help reduce the inequitable burden of anxiety on the Multiracial population. Stochastic methods support consequentialist approaches to racial health equity, and can encourage greater dialogue between public health researchers, policymakers, and practitioners.

## Background

In his influential paper “Sick individuals and sick populations”, Geoffrey Rose argued that epidemiology’s roots in biomedicine often leads to a focus on identifying individual-level risk factors (i.e.: causes of cases), and a failure to recognize the importance of differences in exposure distributions (i.e.: causes of causes, or what Glass and McAtee call risk regulators) across populations in determining disease incidence rates (1,2). This paper initiated a burst of critical public health thinking, some of it focused on how Rose’s “population” strategies – blanket interventions aiming to shift risk distributions of the whole population – might lessen or exacerbate unjust health disparities (i.e. inequities) between groups (3,4). Frohlich and Potvin described an alternative “vulnerable populations” approach, which refocused attention on segments of the population facing excess risk, while reimagining risk as socially (rather than individually) determined (3). More recently developed causal inference methods such as parametric G-computation are useful for epidemiologists interested in moving beyond individual-level risk factors and instead quantifying the differences in population-level risk associated with interventions (5). Stratified causal estimands can answer the question of “who benefits, and by how much?” Such analyses can also identify strong associations in “vulnerable”, small, or oft-ignored populations that may benefit disproportionately from intervention, which is particularly relevant for redressing historical health injustices.

However, causal methods are still infrequently applied in racial health disparities research, where race-stratified analyses or studying race as an effect modifier are more common approaches (6,7). While simple interaction analyses identify whether exposures operate differently in strata of a possible modifier (e.g., race), those working in health disparities are concerned with not just identifying but eliminating disparities. Substitution methods have much to offer to health disparities research because they produce an explicitly consequentialist answer to “who benefits from an intervention, and by how much?” (8–11). Here, we apply these tools in a racial health equity example by studying associations between the high mean ACE scores and prevalence of anxiety among the Multiracial population, a group that is frequently overlooked in population health research but experiences high rates of asthma, anxiety, and depression. Anxiety is characterized by racial and ethnic disparities in prevalence, linked to histories of trauma and other mental illness, and is associated with poor quality of life, substance use, and risk of suicide (12–14). To motivate use of stochastic methods in anxiety disparities research, we begin first with a fictional case study, and then use empirical data from a nationally-representative study to demonstrate the approach.

### Hypothetical scenario

Suppose you are a program planner focused on violence prevention in the Family Health Services Division (FHSD) of the ABC County Department of Health (DoH). ABC County is a very populous, urban, and diverse county in California with a large and growing population of Multiracial people. One day while reviewing literature, you come across a few papers and reports that document stark disparities in both childhood adversity and anxiety between monoracial and Multiracial people (15–20). You know from your work in violence prevention that adverse childhood experiences (ACEs) are strongly associated with anxiety but were unaware of the striking Multiracial-monoracial disparities. You find this data particularly troubling, as you realize that health data on Multiracial people is frequently unavailable, and that Multiracial people are often overlooked in the design of public health interventions (21). Perhaps you are one of the now one in ten Americans that self-identifies as Multiracial and thus have a personal interest in the issue (22).

You reach out to a colleague in the DoH Epidemiology Division and ask for help in pulling some data on the prevalence of ACEs and anxiety, and investigating patterns in their relationship across racial groups. Your colleague asks what your goal is, and you respond, “I want to know 1) if health inequities in anxiety are associated with differential exposure to ACEs, and if so, 2) what racial groups would benefit most if we could eliminate inequities in exposure?” Fortunately, California Senate Bill 428 mandates California health insurance plans to reimburse providers that conduct screening for ACEs, and thus ABC County has a wealth of data on the pediatric prevalence of ACEs (23). Your epidemiologist colleague decides that because you are interested in understanding what populations should be prioritized for intervention, a sensible approach would be to specify a model regressing anxiety on an interaction between ACE scores and race (24). Using this model, you can estimate subgroup-specific risk ratios (RRs) and perform statistical tests to identify which groups experience the strongest associations and should thus be prioritized.

The epidemiologist assembles an appropriate analytic dataset, and runs a model regressing anxiety diagnosis against ACE scores interacted with race, controlling for individual-level characteristics. This model’s results show that ACEs and anxiety are most prevalent for the Multiracial group. However RRs for all racial groups are very close, with narrow and overlapping confidence intervals, and a global Wald test for interaction is insignificant. The epidemiologist concludes that based on the lack of difference in associations by race, there is no evidence that any group should be prioritized for intervention. You present these results to the ABC County health officer and FHSD leadership, who are explicitly interested in health equity and reducing disparities. You suggest that although associations do not seem to vary by race, investing in ACEs prevention could help reduce anxiety inequities for the Multiracial population, given this group’s high exposure to ACEs. The health officer asks you pointedly, “I am not sure results from this analysis support that statement. Show me what kind of reduction in inequity we can expect – I want numbers”. The objective of this paper is to demonstrate one approach to answering this question.

### An alternative approach, using stochastic methods

We present the scenario described here to draw attention to the limitations of statistical interaction analyses in applied health disparities epidemiology. Statistical interactions have useful properties for causal inference in epidemiologic research that are described elsewhere (24). However, they are only one piece of information among many that can help identify appropriate populations for interventions. If we are interested in understanding which populations will benefit most from an intervention, or which populations to intervene on to have the greatest overall impact on reducing racial health disparities, we also need to know the relative sizes of the population subgroups of interest, and the distribution of exposures and associated confounding characteristics across those subgroups.

With these data, we can implement a substitution estimator as an alternative approach to identifying subgroups that will benefit most from a health equity intervention (25). For example, we could estimate and compare the number of cases of disease per 100 that would be averted in each subpopulation if their exposure values were set to approximate that of a relatively more advantaged subgroup, such as those with the lowest exposure levels. We can improve the plausibility of our estimates by introducing some uncertainty in the form of random variation around the substituted exposure value. These “stochastic interventions” (5) specify a distribution of possible exposure values rather than a set deterministic value, which may involve unrealistic assumptions such as universal uptake or perfect adherence (26). Together, these methods provide a principled approach to health disparities analysis that aligns with calls in the field of epidemiology for a stronger consequentialist focus, or a focus on informing policy and programs to improve health (8,27).

We can apply this method to our study question by specifying a regression model similar to the one used by the epidemiologist, and then using it to estimate a population-level metric. These results provide one answer to the question of which subgroups would see the greatest relative reduction in cases from an intervention program to bring health equity in ACEs exposure and anxiety incidence.

## Methods

### Data and analytic sample

To demonstrate the utility of stochastic methods for disparities research, we now move from our hypothetical county to using a nationally-representative dataset with a large sample of Multiracial people for this analysis. The National Longitudinal Study of Adolescent to Adult Health (Add Health), is a prospective cohort of participants followed from adolescence in the 1990’s to early adulthood (28). Eighty public and private high schools and associated feeder schools for each were selected based on specific school characteristics. From these schools’ rosters, 20,745 participants were selected from gender and grade-stratified samples, with oversampling of specific population subgroups to create a representative cohort of all US 7^th^-12^th^ graders in 1994-95. Five waves of interviews taking place from 1994-95 to 2016-18 (response rates ranging from 79-80.3%) collected detailed demographic, life and family history, socioeconomic, educational, health behavior, biomarker, and health and socioeconomic outcomes data. Study design and details are described elsewhere (29–31). We excluded participants self-identifying as “Other” race alone because of numbers too small for meaningful interpretation, and participants self-identifying as Hispanic/Latino ethnicity because of the challenges in identifying Multiracial Hispanic/Latino people when race and Hispanic/Latino ethnicity are assessed separately (32,33).

### Statistical analyses

The primary outcome in this analysis is a binary indicator of diagnosed anxiety at Wave 4. The exposure is a summary ACEs score ranging from 0-10, constructed from questions asked retrospectively across several waves of data collection (see Appendix A for details on score construction). We estimated RRs using a modified Poisson model (34), regressing a binary indicator of diagnosed anxiety at Wave 4 on an interaction between ACEs score on a scale of 0-10 (see Appendix A for details) and Wave 3 race (categorized as monoracial White, Black, Asian, or American Indian/Native American (AI/NA), or Multiracial). Interaction analyses used Whites as the reference as they have a relatively low mean ACE score and large sample size in our data, and are structurally and socioeconomically advantaged compared to other racial groups in the US. We exponentiated interaction term coefficients to evaluate relative interaction, and used the interaction contrast to evaluate additive interaction from our outcome model, estimating excess cases per 1,000 (35). The outcome model specified the following covariates to be confounders and controlled for them using data from the indicated waves: Wave 4: age, self-reported gender; Wave 1: highest parental education (categorized ordinally as less than high school, completed vocational school, or equivalency (GED); high school diploma (HSD); some college; college graduate or greater), household size-adjusted income (continuous equivalence scale of pretax household income divided by the square of household size), parental support (mean of five Likert-scale questions about warmth, communication, caring, and closeness in relationship with parents), and neighborhood disadvantage score (mean of census-tract proportions of households earning less than federal poverty level, households receiving public assistance, civilian unemployment, persons 25 years or older lacking HSD or GED, and female-headed households). Further details on variable construction are available in Appendix B.

### Stochastic intervention

Our target causal quantity is the population mean risk difference (*RD_r_*) of anxiety (*Y*) if each non-White racial group’s (*R = r*) distribution of ACEs (*A*) reflected the exposure distribution of the White (*R = w*) population given an individual’s covariates (*X*). We chose the ACEs distribution of Whites because this group on average has a low mean ACE score and the largest sample size to provide support for precision in estimates of risk differences. Let *g_w_* denote the ACEs distribution for Whites conditional on covariates *X*. We estimate this parameter following an approach similar to that outlined in Ahern et al (5). We can then represent the target quantity as a risk difference using the following notation, where inner expectations are over the exposure distribution given race, and outer expectation over covariates to estimate population-level average risk:

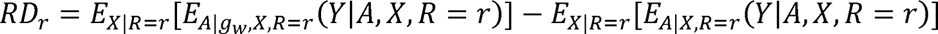

To estimate this distribution, we used Monte Carlo simulation methods to represent interventions on the ACE score for non-White participants with some stochastic variation (5,36). Specifying a substitution estimator that accounts for the stochastic nature of the intervention across the population, rather than intervening on ACE scores with a static, uniform value, is appealing as it is often unrealistic to assume that every person receiving the intervention is affected in a uniform manner (5,36). Specifying a substitution estimator with variation in the intervention is beneficial as it is unrealistic to assume that every person receiving the ACEs prevention intervention would with certainty experience the targeted (mean of White group) ACEs score (26). To estimate the number of cases of anxiety averted for each racial group, we average over *i* = 1,000 simulated risk differences for each racial group. To calculate simulation-based confidence intervals (here forth CIs), we use parameter resampling instead of bootstrapping (37–39) as it is more computationally efficient for complex estimands such as population-level parameters that incorporate survey design effects.

We used complex survey weights to incorporate design effects using the “survey” package in R (40). Because of high levels of missingness in ACE components, covariates, and outcomes (distributions and frequencies of missing information available in Appendix C), we performed multiple imputation to reduce biased estimation due to missing data using the “mi” and “mitools” packages in R (41,42). We performed Markov chain imputation with 30 iterations and 4 chains, including the anxiety outcome in our imputation model, and pooled results across 20 imputed datasets.

The following steps summarize implementation of the stochastic intervention approach, which is based off the approach of Ahern et al (5).

1. First, we use the estimated beta coefficients and variance-covariance matrix from our outcome model to generate a single resampled value of the beta coefficients from a multivariate normal distribution (37–39).
2. We then resample covariate values with replacement *m* times from one imputed data frame, where *m* = number of observations in the dataset. We use these resampled data to get the distribution of covariates under no intervention.
3. We use the coefficients sampled in step 1 to obtain the predicted probability of anxiety for each observation, and then sum across all observations for each racial group to estimate the race-specific prevalence of anxiety under no intervention.
4. Next, we create a copy of the resampled data from step 2, replacing the ACEs score for all non-White participants with simulated value from a Poisson distribution, with the rate parameter estimated from a Poisson model fit among White participants.
5. We repeat step 3 to calculate the prevalence of anxiety in this single draw from the intervention distribution.
6. We calculate the risk differences by race as the averaged risk differences across each observation in the various racial groups from the resampled population.
7. Finally, we repeat this procedure 1,000 times to build up the sampling distribution of the race-specific RD. From these 1,000 resampled estimates, we report median differences as race-specific point estimates and quantile-based 95% CIs around the risk differences.

Example code in R for this simulation is provided as a link in this article’s supplementary material.

## Results

Table 1 summarizes parameters from the outcome model. Mean ACE scores were highest among AI/NA (3.16), Multiracial (2.90), and Black (2.84) groups. Anxiety prevalence was highest among the Multiracial (18.2%) and White (15.0%) groups. An additional ACE was weakly associated with increased risk of anxiety among White (RR = 1.02, 95% CI: 1.01, 1.02) and Multiracial (RR = 1.03, 95% CI: 1.02, 1.05) groups. RRs for other groups were centered around the null, with narrow CIs. The models predicted relatively fewer excess cases per 1,000 for Black (−7.94), Asian (−20.2), and AI/NA (−18.5) participants compared to Whites, but more for Multiracial (17.0) participants. Confidence intervals were most precise around estimates for the Black group (95% CI: −18.1, 1.88), and least precise for the AI/NA estimate (95% CI: −41.4, 1.93).

Table 2 presents the baseline and intervened ACE scores for each racial group, and a summary of the population-averaged results of our simulated intervention. Mean resampled baseline ACE scores are slightly lower than in the original dataset, reflecting the influence of outliers in the original data which were less frequently selected in repeated resampling. Differences in mean baseline scores are most pronounced for Black participants (original = 2.84, resampled = 2.69). Figure 1 shows that our stochastic intervention shifted the ACEs distribution lower most substantially for the AI/NA population, followed by substantial shifts in distribution for the Black and Multiracial populations. For the Asian population, the distribution shift was mixed, and resulted in fewer individuals with no ACEs and more individuals with ACE scores of 2 and 3. The intervention resulted in a slightly higher mean ACE score for the Asian group; we report the means here for consistency of presentation, but a real intervention would never be designed to increase one group’s exposure to a putative risk factor. We did not intervene on the White distribution, and so there was no change in shape for that group.

The median risk difference was largest for the Multiracial population, with 4.17 cases per 1,000 averted under repeated draws of the simulation. Our model estimated that the intervention would prevent 0.76 cases per 1,000 among the Black population. The model predicted an decrease of 0.01 cases per 1,000 for Asians, and an increase of 0.03 cases per 1,000 for AI/NA participants. CIs for the Asian and AI/NA groups both crossed the null, and were wider for the AI/NA group..

Figure 2 plots the population-averaged risk differences produced in each of our 1,000 intervention draws across 1,000 resampled datasets. Estimate precision was greatest for Black, Asian, and Multiracial groups; there was much higher variability in the risk differences for AI/NA groups. These are reflected in the quantile-based 95% CIs in Table 2.

**Table 1.** Summary of results from adjusted^a^ interaction models for anxiety, Add Health 1994-2008

| Group | n (%) | Mean score<sup>b</sup> | Anxiety prevalence<sup>c</sup> | Adjusted RR<sup>d</sup><br>(95% CI) | Excess cases<sup>e</sup> per<br>1,000 (95% CI) |
| --- | --- | --- | --- | --- | --- |
| White | 7,742 (73.5%) | 2.34 | 15.2% | 1.02 (1.01, 1.02) | (ref.) |
| Black | 2,915 (16.9%) | 2.84 | 5.95% | 1.01 (1.00, 1.02) | -7.94 (-18.1, 1.88) |
| Asian | 805 (3.18%) | 2.30 | 2.93% | 1.00 (0.99, 1.01) | -20.2 (-31.4, -10.0) |
| AI/NA | 76 (0.55%) | 3.16 | 3.26% | 1.00 (0.98, 1.02) | -18.5 (-41.4, 1.93) |
| Multiracial | 834 (5.28%) | 2.90 | 17.6% | 1.03 (1.02, 1.05) | 17.0 (-1.15, 34.3) |

<sup>a</sup> Outcome model adjusted for participant age, sex, parental education, household size-adjusted income, parental support, and neighborhood disadvantage score
<sup>b</sup> Individual weighted scores summed after imputation of missing ACE components
<sup>c</sup> Weighted prevalence
<sup>d</sup> RR associated with an additional ACE, within racial group
<sup>e</sup> Excess cases estimated using interaction contrast (additive excess risk)

**Table 2.**
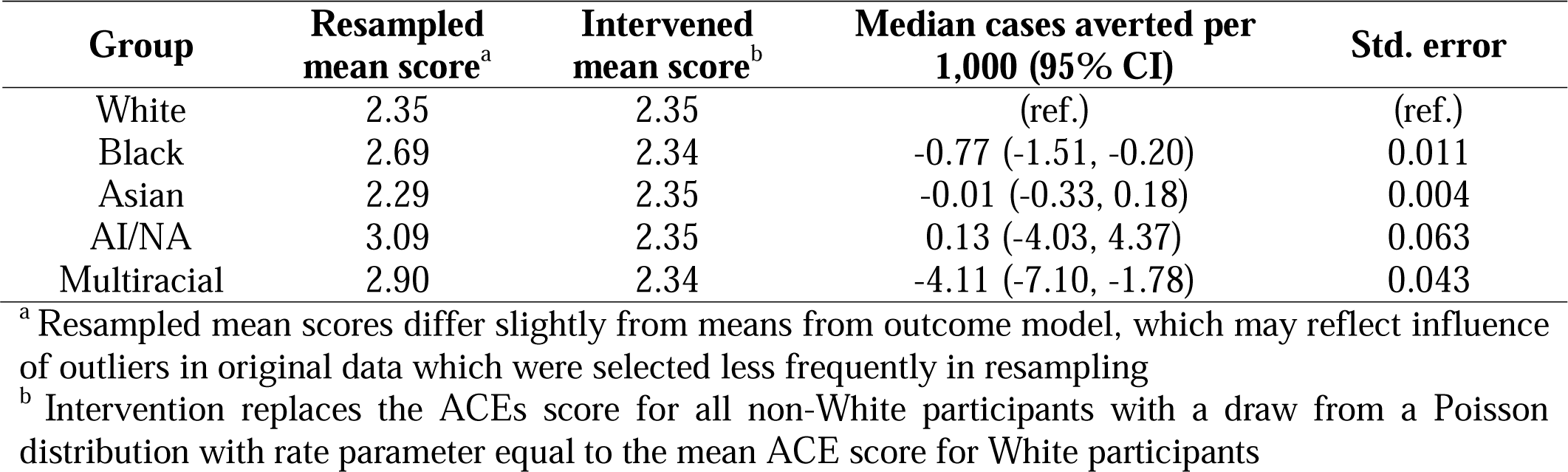
Summary of mean ACE scores and cases of anxiety averted per 1,000 under stochastic intervention, Add Health 1994-2008

**Figure 1.**
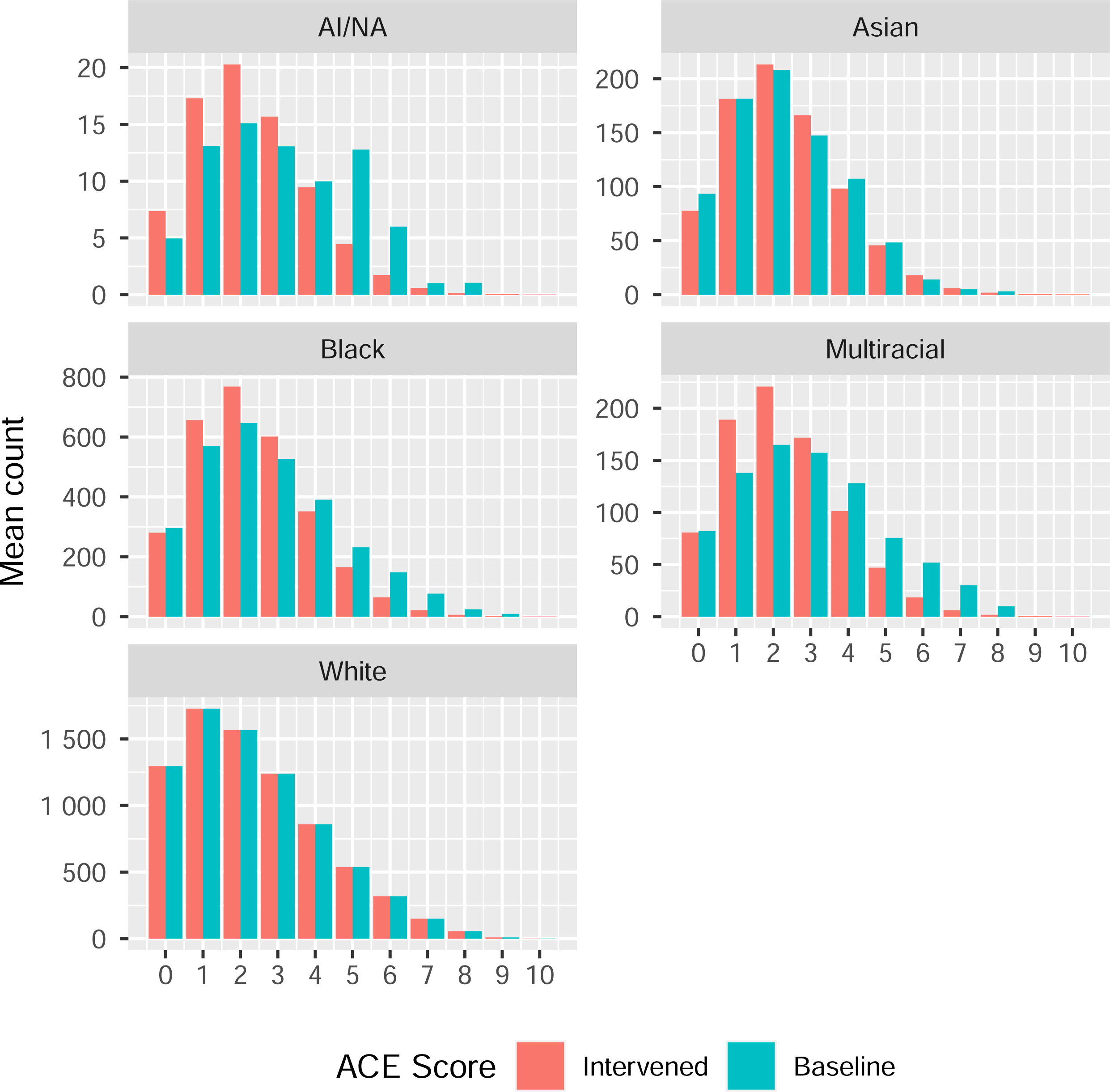
Comparison of intervention and baseline ACE score distributions resulting from 1,000 resampled datasets, Add Health 1994-2008

**Figure 2.**
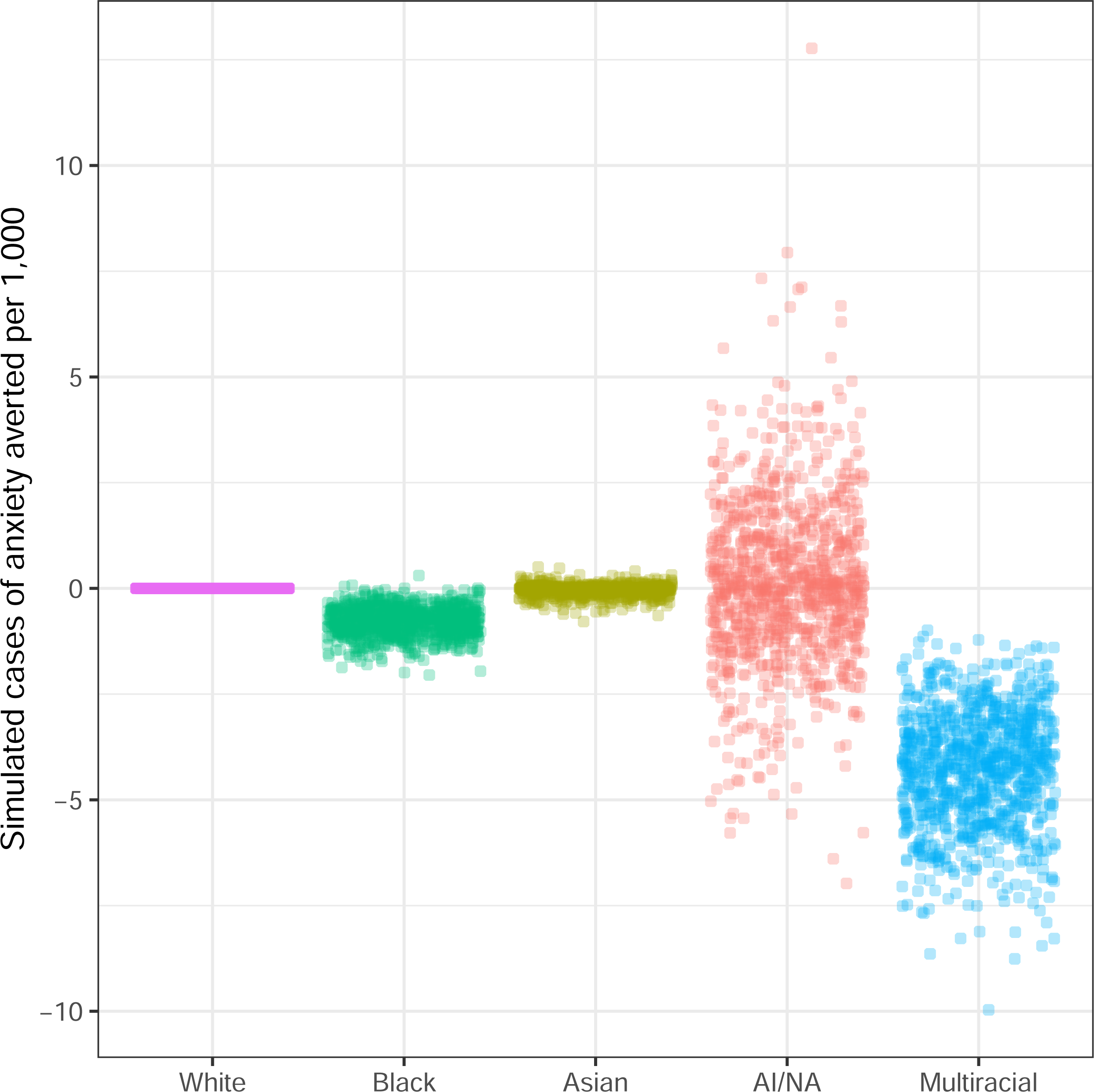
Simulated number of cases of anxiety averted per 1,000 population, results from 1,000 resampled datasets, Add Health 1994-2008

## Discussion

In this paper we illustrate a principled approach to estimating differences in the population impact of a hypothetical intervention in defined subgroups, as one way of identifying groups that might benefit the most from the intervention. Our analysis shows that there are substantial racial differences in the number of anxiety cases averted associated with an intervention to reduce disparities in exposure to ACEs, with the Multiracial population standing to benefit most from such an intervention. These differences are easily masked in subgroup analyses, where population sizes and distribution of exposures are not usually considered. However, as we have demonstrated, stochastic intervention models allow us to quantify the reduction in burden of disease across racial groups at a population level under altered exposure distributions.

Ahern et al (5) position stochastic methods as a way to study the potential effects of population- or community-level policies on exposure distributions and their associations with changes in incidence of disease. However, these methods are also a natural fit for consequentialist health equity research. Health equity exists when everyone has a fair and equal opportunity to realize their full health potential, which requires addressing inequity in exposure to risk factors for disease (43). Modeling shifts in exposure distributions – or elimination altogether of disparities – allows us to both envision and quantify a more equitable counterfactual reality. Furthermore, stochastic methods (and other modern approaches such as G-methods) (44,45) do not require deterministically setting counterfactual exposure values, providing analytic flexibility not offered by more traditional regression and standardization approaches. Finally, stochastic methods provide an alternative approach to identifying priority subgroups for intervention, which is most frequently assessed using interaction (and increasingly, mediation) (46) analyses.

A natural next step for practitioners interested in reducing anxiety incidence among the Multiracial population would be to contextualize the results from this simulation with other potential interventions, and potentially to compare costs in a cost-effectiveness analysis. These methods can be extended to model different kinds of interventions – for example, a uniform reduction in the ACEs score rather than a shifting of distributions – or an intervention reducing the ACE score of individuals within a given strata of socioeconomic position. Our alteration of exposure patterns is just one example of a potential intervention to reduce inequities in anxiety, but it draws attention to the differential impact by racial group such an intervention could produce.

This study had limitations. Our estimates can only be interpreted as causal under the identifiability assumptions: exchangeability, stability, positivity, and consistency (47). Modeling the exposure as a count of ACEs may challenge positivity given the relative rarity of participants reporting 6 or more ACEs. High variability in the AI/NA group’s RR estimates from the underlying outcome model are not eliminated with repeated resampling; future studies should make greater efforts to include larger proportions of vulnerable populations to improve inference. Our demonstration is purely hypothetical because it models intervening directly on ACEs; in reality ACE prevention strategies target determinants of ACEs such as family psychosocial or economic stress or caring adult presence (48). Simulating interventions to modify *determinants* of ACEs would have more policy relevance and thus better align with calls for consequentialist epidemiologic research. Still, we believe simulating intervention on ACE scores is useful for demonstrating the limitations of focusing solely on identifying subgroup associations. Finally, our simulations do not specify a dependence between the intervened ACEs score and an individual’s set of covariates. It is unlikely that the effect of a real intervention would be random; instead, effects would likely be associated with an individual’s SEP, environment, access to care, and other characteristics. Incorporating such individual-level covariate data to model a real-world intervention more realistically would improve quality of inference.

The Multiracial population’s disproportionate exposure to ACEs and prevalence of anxiety are concerning, yet this group is rarely highlighted in discussions of health equity, let alone prioritized for receiving preventative interventions. Our analysis shows that racial inequities in exposure to ACEs are associated with substantial population-level disparities in anxiety, and that the Multiracial population would benefit significantly from a reduction in ACEs compared to other groups. Strategies to address inequities in anxiety for the Multiracial population may benefit by including interventions to reducing exposure to ACEs among Multiracial children and families. Public health researchers and practitioners should employ rigorous methodologies to weigh different interventions and suitable target populations to remedy inequities in exposures and health outcomes. Health equity will only be achieved when disparities between populations are addressed, and stochastic methods are one among many that can help us make progress towards those aims.

## Conflict of interest statement

This work was supported by NIH-NCATS-CTSA grant UL1TR003142 and contract 75D30122P12974 with the Centers for Disease Control and Prevention.

## Financial disclosures

No financial disclosures were reported by the authors of this paper.

## Data Availability

This analysis used restricted data (made available through a data use agreement); a public-use subset is available online.

https://addhealth.cpc.unc.edu/data/

## Acknowledgements

We would like to thank Add Health study participants for their time, energy, and information which provide invaluable contributions to science. Thanks as well to Kim Harley for providing access to the data, and to Christian Jackson for reviewing the code used in this analysis.

## Data and code availability

This analysis used restricted data (made available through a data use agreement); a public-use subset is available at https://addhealth.cpc.unc.edu/data/. Example code in R for this analysis is available at https://github.com/lamhine/stochastic_intervention.

## Notes

### Competing Interest Statement

The authors have declared no competing interest.

### Clinical Protocols

https://github.com/lamhine/stochastic_intervention

### Author Declarations

The Office for Protection of Human Subjects of the University of California Berkeley waived ethical approval for this work

