## Appendices for "A hypothetical intervention to reduce inequities in anxiety for Multiracial people: simulating an intervention on childhood adversity"

**Appendix A.** Crosswalk of topical domains, questions in Kaiser ACEs questionnaire, and questions in Add Health used to create ACEs score

| **Domain** | **ACE Questionnaire** | **Add Health Measure Coding** |
| --- | --- | --- |
| Emotional  abuse | 1. Did a parent or other adult in  the household often...  Swear at you, insult you, put you down, or humiliate you?  or  Act in a way that made you afraid that you might by physically hurt?  Yes/No | H4MA1 (Wave 4): Before your 18th birthday, how often did a parent or other adult caregiver say things that really hurt your feelings or made you feel like you were not wanted or loved?  1 = 1-10 times or more  0 = this never happened |
| Physical  abuse | 2. Did a parent or other adult in  the household often...  Push, grab, slap, or throw some- thing at you?  or  Ever hit you so hard that you had marks or were injured?  Yes/No | H3MA3 (Wave 3): Before the time you started 6th grade, how often had your parents or other adult care- givers slapped, hit, or kicked you?  1 = 1-10 times or more  0 = this never happened |
| Sexual abuse | 3. Did an adult or person at least 5 years older than you ever... Touch or fondle you or have you touch their body in a sexual way?  or  Try to actually have oral, anal, or vaginal sex with you?  Yes/No | H3MA4 (Wave 3): Before the time you started 6th grade, how often had your parents or other adult care- givers touched you in a sexual way, forced you to touch him or her in a sexual way, or forced you to have sexual relations?  1 = 1-10 times or more  0 = this never happened |
| Emotional  neglect | 4. Did you often feel that...  No one in your family loved you or thought you were important or special?  or  Your family didn’t look out for each other, feel close to each other, or support each other?  Yes/No | H1PR8 (Wave 1): How much do you think your family pays attention to you?  1 = quite a bit - very much  0 = not at all - somewhat |
| Physical  neglect | 5. Did you often feel that...  You didn’t have enough to eat, had to wear dirty clothes, and had no one to protect you?  or  Your parents were too drunk or high to take care of you or take you to the doctor if you needed it?  Yes/No | By the time you started 6th grade, how often had your parents or other adult care-givers:  H3MA1 (Wave 3): Left you home alone when an adult should have been with you?  or  H3MA2 (Wave 3): Not taken care of your basic needs, such as keeping you clean or providing food or clothing?  1 = 1-10 times or more for either  0 = this never happened |
| Parental separation or divorce | 6. Were your parents ever separated or divorced?  Yes/No | PA38 - PA54 (Wave 1 Parent)  1 = parents divorced or separated before participant turned 18  0 = parents not divorced or separated before participant turned 18 |
| Mother  treated violently | 7. Was your mother or step-mother:  Often pushed, grabbed, slapped, or had something thrown at her?  or  Sometimes or often kicked, bitten, hit with a fist, or hit with something hard?  or  Ever repeatedly hit over at least a few minutes  or  Threatened with a gun or knife?  Yes/No | PB20 (Wave 1 Parent): How much do you fight or argue with your current (spouse/partner)?  1 = a lot  0 = not at all - some |
| Household  substance abuse | 8. Did you live with anyone who  was a problem drinker or alcoholic or who used street drugs?  Yes/No | PC49E_2 (Wave 1 Parent): His/her biological father has alcoholism?  or  PC49E_3 (Wave 1 Parent): His/her biological mother has alcoholism?  or  H1TO52 (Wave 1): Are illegal drugs easily available to you in your home?  1 = yes to any  0 = no to all |
| Mental illness  in household | 9. Was a household member depressed or mentally ill or did a household member attempt suicide?  Yes/No | PA20 (Wave 1 Parent): In general,  are you (main parent respondent) happy?  or  PB16 (Wave 1 Parent): In general do you think (he/she) (main parent respondent’s partner or spouse) is happy?  or  H1SU6 (Wave 1): Have any of your family tried to kill themselves during the past 12 months?  1 = yes to PA20, yes to PB16, and no to H1SU6  0 = any other combination of responses |
| Incarceration of household member | 10. Did a household member go to  prison?  Yes/No | H4WP3 (Wave 4): Has/did your biological mother ever spent/spent time in jail or prison?  or  H4WP9 (Wave 4): Has/did your biological father ever spent/spend time in jail or prison?  or  H4WP16 (Wave 4): Has/did your mother figure ever spent/spend time in jail or prison?  or  H4WP30 (Wave 4): Has/did your father figure ever spent/spend time in jail or prison?  1 = yes to any  0 = no to all |

**Appendix B.** Key questions used to construct Add Health covariate and outcome measures

| **Variable** | **Add Health Questions** | **Coding** |
| --- | --- | --- |
| Metabolic  syndrome | Wave 4 and Biomarker File  Hypertension  Systolic blood pressure ≥ 130 mmHg (H4SBP), or  Diastolic blood pressure ≥ 80 mmHg (H4DBP), or  Ever diagnosed with hypertension (H4ID5C), or  Current hypertension medication use (medication file)  Waist circumference (> 102 cm for males, > 80 cm for females) (H4WAIST)  Triglycerides (top 3 deciles for males, top 2 for females)  High-density lipoprotein (bottom 2 deciles for males, bottom 3 for females)  Glycated hemoglobin (HbA1c) > 5.7%. | 1 = exceeding cutoff in three or more of the five categories  0 = exceeding cutoff in fewer than three categories |
| Asthma | H4ID5F (Wave 4)  Has a doctor, nurse or other health care provider ever told you that you have or had asthma, chronic bronchitis or emphysema?  Yes/No | 1 = yes  0 = no |
| Depression | H4ID5H, H4MH18-27 (Wave 4)  Have you taken prescription medication in the past 12 months for depression or stress  or  Has a doctor, nurse or other health care provider ever told you that you have or had depression?  Yes/No | 1 = yes to either  0 = no to both |
| Anxiety | H4ID5J (Wave 4)  Has a doctor, nurse or other health care provider ever told you that you have or had: anxiety or panic dis- order?  Yes/No | 1 = yes  0 = no |
| Suicidal  ideation | H4SE1 (Wave 4)  During the past 12 months, have you ever seriously thought about committing suicide?  Yes/No | 1 = yes  0 = no |
| Problematic  drug use | H4TO63, H4TO65C-E, H4TO66 (Wave 4)  Have you ever taken any prescription drugs that were not prescribed for you, taken prescription drugs in larger amounts than prescribed, more often than pre- scribed, for longer periods than prescribed, or taken prescription drugs that you took only for the feeling or experience they caused?  or  Have you ever used cocaine, crystal meth, or other drugs (not including marijuana or steroids)?  or  Have you ever injected (shot up with a needle) any illegal drug, such as heroin or cocaine?  Yes/No | 1 = yes to any  0 = no to all |
| Parental  support | H1PF1, H1PF4, H1PF5, H1NM14, H1WP10,  H1PF23-25, HI1NF14, H1WP14 (Wave 1)  Most of the time, your mother/father is warm and loving toward you  You are satisfied with the way your mother/father and you communicate with each other  Overall, you are satisfied with your relationship with your mother/father  How close do you feel to your mother/father?  How much do you think your mother/father cares about you?  Very much, quite a bit, some, a little, none at all) | 5 = very much  4 = quite a bit  3 = some  2 = a little  1 = none at all |
| Racial discrimination | H4MH29 (Wave 4)  What do you think was the main reason for being treated with less respect or courtesy than other people?  National ancestry or origin, gender, race, age, religion, height or weight, shade of skin color, sexual orientation, education or income, physical disability, other | 1 = national ancestry or origin, race, or shade of skin color  0 = all others |

**Appendix C.** Participant characteristics^a^ comparing complete case^b^, imputations^c^, and missingness^d^, stratified by race

| **Characteristic** | **White** | **Black** | **Asian** | **AI/NA** | **Multiracial** | **Overall** |
| --- | --- | --- | --- | --- | --- | --- |
|  | 7,742 (73.5%) | 2,915 (16.9%) | 805 (3.18%) | 76 (0.55%) | 834 (5.82%) | 12,372 (100%) |
| Male sex |  |  |  |  |  |  |
| Complete case | 3,672 (51%) | 1,267 (50%) | 418 (53%) | 41 (64%) | 380 (50%) | 5,778 (51%) |
| Imputed | 51% | 50% | 53% | 41% | 50% | 51% |
| Missing | 0 (0%) | 0 (0%) | 0 (0%) | 0 (0%) | 0 (0%) | 0 (0%) |
| Age |  |  |  |  |  |  |
| Complete case | 28.9 | 29.2 | 29.2 | 28.8 | 28.8 | 29.0 |
| Imputed | 28.9 | 29.2 | 29.2 | 28.8 | 28.8 | 29.0 |
| Missing | 0 (0%) | 0 (0%) | 0 (0%) | 0 (0%) | 0 (0%) | 0 (0%) |
| Highest parental education |  |  |  |  |  |  |
| Less than high school^e^ |  |  |  |  |  |  |
| Complete case | 685 (9.6%) | 356 (17%) | 69 (13%) | 13 (29%) | 64 (11%) | 1,187 (11%) |
| Imputed | 9.6% | 17% | 13% | 29% | 11% | 11% |
| High school diploma |  |  |  |  |  |  |
| Complete case | 2,520 (34%) | 959 (42%) | 164 (24%) | 19 (34%) | 221 (33%) | 3,883 (35%) |
| Imputed | 34% | 42% | 23% | 34% | 33% | 35% |
| Some college |  |  |  |  |  |  |
| Complete case | 1,405 (19%) | 500 (16%) | 106 (10%) | 19 (24%) | 191 (22%) | 2,221 (18%) |
| Imputed | 19% | 16% | 10% | 24% | 22% | 18% |
| College graduate or greater |  |  |  |  |  |  |
| Complete case | 3,039 (38%) | 1,043 (25%) | 445 (53%) | 24 (14%) | 346 (34%) | 4,897 (36%) |
| Imputed | 38% | 26% | 53% | 14% | 34% | 36% |
| Missing | 93 (1.4%) | 57 (2.5%) | 21 (2.0%) | 1 (<0.1%) | 12 (1.5%) | 184 (1.6%) |
| Household income^f^ |  |  |  |  |  |  |
| Complete case | 3.46 | 2.03 | 3.89 | 1.56 | 2.78 | 3.21 |
| Imputed | 3.56 | 2.12 | 3.85 | 1.96 | 2.89 | 3.28 |
| Missing | 1,481 (19%) | 850 (30%) | 338 (40%) | 24 (27%) | 186 (20%) | 2,879 (21%) |
| Parental support index (0-5) |  |  |  |  |  |  |
| Complete case | 3.35 | 3.26 | 3.30 | 3.39 | 3.30 | 3.33 |
| Imputed | 3.35 | 3.26 | 3.30 | 3.38 | 3.30 | 3.33 |
| Missing | 9 (0.2%) | 9 (0.3%) | 2 (0.3%) | 1 (1.0%) | 0 (0%) | 21 (0.2%) |
| Neighborhood disadvantage (0-1) |  |  |  |  |  |  |
| Complete case | 0.12 | 0.23 | 0.13 | 0.23 | 0.15 | 0.14 |
| Imputed | 0.12 | 0.23 | 0.13 | 0.23 | 0.15 | 0.14 |
| Missing | 80 (1.2%) | 28 (1.1%) | 4 (0.6%) | 0 (0%) | 9 (1.1%) | 121 (1.1%) |
| ACEs^g^ |  |  |  |  |  |  |
| Complete case | 1.89 | 2.12 | 1.92 | 2.58 | 2.39 | 1.94 |
| Imputed | 2.34 | 2.84 | 2.30 | 3.16 | 2.91 | 2.46 |
| Missing | 3,899 (50%) | 2,127 (75%) | 499 (59%) | 55 (79%) | 481 (54%) | 7,061 (55%) |
| Anxiety |  |  |  |  |  |  |
| Complete case | 1,179 (15%) | 182 (6.0%) | 26 (2.9%) | 4 (3.3%) | 127 (18%) | 1,518 (13%) |
| Imputed | 15% | 6.0% | 2.9% | 3.3% | 18% | 13% |
| Missing | 2 (<0.1%) | 0 (0%) | 0 (0%) | 0 (0%) | 0 (0%) | 2 (<0.1%) |

Abbreviations: ACE = adverse childhood experience; RERI = relative excess risk due to interaction

^a^ Unweighted counts

^b^ Weighted proportions (for categorical variables) and means (for continuous) reported

^c^ Imputations pooled over 20 datasets; imputation models included all regression variables and variables representing status of parental self-rated health, divorce, employment, disability, retirement, happiness, and welfare receipt, interviewer assessments of neighborhood safety and how well-kept the household is, and number of interruptions to interview with parent

^d^ Unweighted counts and weighted proportions

^e^ Includes completed vocational school or GED

^f^ Equivalence-scaled to adjust for household size

^g^ ACE components measured variously across Waves 1, 3, and 4, see Appendix A for details
